# Healthcare providers’ knowledge and use of psychological and psychoSOCIAL screening and interventions in the management of patients with tendinopathy: An International Survey of Practice

**DOI:** 10.1101/2024.06.05.24308397

**Authors:** Seth O’Neill, Laura English, Adrian Mallows, Carl Stubbs, Gareth Stephens, Sam Briggs-Price, Paul Kirwan, Matt Lee, Sean McAuliffe, Matt Kenyon

## Abstract

**Background:** Psychological and psychosocial factors play an important role in the management of patients with musculoskeletal disorders. Currently, there is no information exploring how clinicians current practice is informed by these factors in the people with tendinopathy exists.

**Objectives:** To explore healthcare providers knowledge and use of psychological and social screening and interventions in the management of patients with tendinopathy

**Methods:** An online survey was developed by a group of tendon experts. The survey was disseminated via special interest groups, social networks and professional organisations internationally.

**Results:** The survey had 103 completed responses. The majority of respondents used the subjective, 53% (N=55), subjective and objective, 28%(n=29), or objective 6% (n=6) examination to screen for psychological factors using both verbal and nonverbal methods. Psychosocial factors were screened for during the subjective assessment by 75% (n=77) of respondents. A further 15% (n=15) screened during the subjective and objective combined whilst 5% (n=5) examined this factor in the objective assessment in isolation.

Psychological screening tools were used by 25% (n= 26) of respondents and psychosocial factors by 12% (n=12) of respondents.

Treatment typically comprised of individualised education, reassurance, addressing mal-adaptive behaviours and behaviour change. Confidence in assessment and treatment was mixed and clinicians identified a desire for more specific training and self-development.

**Conclusion:** The proportion of clinicians screening and measuring psychological and psychosocial factors in clinical practice is high, but few use validated tools due to a lack of time and confidence.

**Implication for clinical practice:** Clinicians commonly assess psychological and social factors during assessment of individuals with tendinopathy, as part of their subjective and objective assessments. It is unclear how successfully clinicians identify these factors during their assessments, as they rarely use validated screening tools.

**Key message –:**

- Clinicians and researchers should examine and modify: fear of movement, Negative pain beliefs, Maladaptive/avoidance behaviors, catastrophisation and Anxiety (psychological constructs) and Quality of life, work related constructs, sleep quality, education health literacy and social interactions(psychoSOCIAL constructs) during clinical or research work.
- Training needs to be developed to improve clinician confidence when assessing and treating psychological and psychoSOCIAL factors in patients with tendinopathy
- Further work is needed to examine the barriers and facilitators to the use of appropriate validated psychological and psychoSOCIAL tools in clinical care.

## Background

Musculoskeletal-related conditions are the largest long-term health condition, affecting 32% of the population and costing £5 Billion annually in the UK alone. [1] Lower limb tendon conditions are now recognised to be more common than lower limb osteoarthritis (OA), [2] but despite this a persistent discrepancy exists in healthcare training and literature. Lower limb tendon disorders impact quality of life to a similar, or greater degree than OA. [3–6] However, the extent to which these factors are screened and managed in clinical practice remains unclear, leading to less effective current management solutions. This contributes to 20% of patients with Achilles tendinopathy and 45% of gluteal tendinopathy patients still having pain at 10 years after initial onset. [7,8] Furthermore, gluteal tendinopathy increases the risk of developing hip OA by 21 times. [7] The long-term consequences of ongoing pain and reductions on physical activity levels further compound the problem and are thought to contribute to secondary co-morbidities and the significant economic costs. [1,7]

Psychological and psychoSOCIAL factors play an important role in the management of patients with musculoskeletal disorders. [8,9] In recent years we have seen a greater degree of interest around psychological and psychoSOCIAL factors in relation to tendinopathy. [10–18] This has led to the identification of specific subgroups who have a psychologically:psychoSOCIALLY dominant profile. [19–21] Follow on studies have identified divergent clinical outcomes for sub-groups of people with Achilles tendinopathy. [22] Notably, the psychologically dominant phenotype appears to exhibit less favourable responses to usual care, suggesting the psychologically dominant subgroup, and potentially all subgroups, could benefit from more tailored treatment encompassing a truly holistic rehabilitation programme.

The recent International Scientific Tendon Symposium ICON 2020 papers highlight the large variations in outcome measures [12] and specifically psychological and psychoSOCIAL measures used. [13] The recent Delphi study explored which psychological and psychoSOCIAL constructs should be used by researchers and clinicians dealing with patients with tendinopathy. [10] Whilst these data are useful, it is important we understand the clinical utilization of such tools and constructs. As such, we decided to complete an international survey of healthcare clinicians to assess their utilization of psychological and psychoSOCIAL factors in patients with tendinopathy so that this information may guide implementation research.

### Aim

To explore healthcare providers’ knowledge and use of psychological and psychoSOCIAL screening and interventions in the management of patients with tendinopathy

## Methods

An online survey was developed by a group of tendon experts (n=7, SON, AM, CS, GS, PK, SMc, and MK) involved in the ICON scoping review and follow up Delphi study examining Psychological and PsychoSOCIAL constructs in tendinopathy. We based the tool on a previous national survey [23] we had completed and modified accordingly. The tool was refined through eight rounds with the tendon experts and transferred into JISC online surveys platform for dissemination. The survey consisted of questions related to participants demographics with a focus on clinical background (healthcare profession, time in profession, area of clinical work, post graduation qualifications) alongside information focused on psychological and psychoSOCIAL constructs (do you assess these components, when are these assessed and how are these assessed) later questions then asked participants to rank pre-populated constructs in order. These were based on the previous scoping review we were involved in. It was possible for additional constructs to be added throughout the survey. Finally, the confidence of respondents’ examination of psychological and psychoSOCIAL assessment was examined. The survey used both open and closed questions.

Ethical approval was sought and granted from the University of Leicester Research Ethics Committee (31603-is489-ls:medicine).

### Dissemination and data analysis

The survey was disseminated via an electronic link allowing direct access to the survey. Paper surveys were not used, which may influence responses to those who are digitally active, however this method also reduces the likelihood of inputting errors and costs. Dissemination pathway included special interest groups (AFAP, MACP) and social media platforms and networks (Twitter/X, Facebook, email lists) and professional organisations (iCSP) and from participants directly to work colleagues on local circulation lists.

Data analysis was descriptive and used the embedded tools in JISC online surveys, Microsoft Excel and SPSS version28. WordClouds were used to report the open questions.

### Inclusion criteria

The survey was open to any practicing healthcare professional with self-reported experience of treating people with tendinopathies (upper and/or lower limb). Students and those with no experience of treating people with tendinopathy were excluded. This criterion was set to be as inclusive as possible and to ensure the full spectrum of healthcare professions involved in patient care was included. Incomplete surveys were excluded.

## Results

### Response to the Survey

A total of 103 responses were collected during the 12 weeks the survey ran. 79 of these respondents were Physiotherapists (76.7%) and there were 4 surgeons (3.9%), 5 physicians (4.9%), 5 clinical academics (4.9%), 3 sports therapists (2.9%), 1 osteopath (1%), 2 chiropractors (1.9%), 2 GPs (1.9%) and 2 “others” (1.9%). The “others” were 1 surgical care practitioner and 1 podiatrist. The survey was opened 1341 times and there were 142 incomplete responses. Out of the 103 respondents, 66 (64.1%) were from the UK, 9 participants (8.7%) were from Australia, 6 participants (5.8%) were from Ireland, 11 participants (10.7%) were from Europe, 4 participants (3.9%) were from the USA, and 7 participants (6.8%) selected other. People selecting “other” worked in Turkey (N=1), Brazil (N=1), Ecuador (N=1), Canada (N=2) and India (N=2).

### Clinical setting

The work settings varied with many of the respondents working in multiple locations. The majority worked in a public hospital, 22 participants (21.4%), whilst a further 15 (14.6%) worked in a public hospital and elsewhere. Nine (8.7%) participants worked solely in private hospitals and 16 (15.8%) worked in community/public clinics, whilst 20 (18.7%) worked in private therapy centers or clinics. The remaining 23 (22.3%) participants worked across professional sports in isolation (N=9, 8.7%) or combined professional sports with academic roles.

### Experience and interest

83 (81.6%) of the respondents have 6 years or more clinical experience, 30 (29.1%)) have 21+ years experience. 87 (84.5%) of the respondents see 6 or more patients a month with a tendinopathy, 66 (64.1%) of them see 11 or more patients a month. 53 (51.5%) respondents highlighted that they have a specialist interest in tendinopathy.

### Assessment of Psychological and PsychoSOCIAL Constructs

Participants were asked in what area of their assessment they assess psychological and psychoSOCIAL factors. Specifically, we were seeking whether these factors were assessed as a part of a verbal discussion (subjective assessment), the physical assessment (objective assessment) or with a ‘Specific Screening Tool’.

### Assessment of Psychological Components

#### Subjective Assessment

The majority of respondents, 55 (53.4%), said that they screen for psychological factors during their subjective assessment with their patient and 29 (28.2%) screened using the subjective and objective assessments. Respondents who selected subjective assessment were then asked to explain how they do this. Respondents answers are represented in a word cloud, Figure 1.

**Figure 1.**
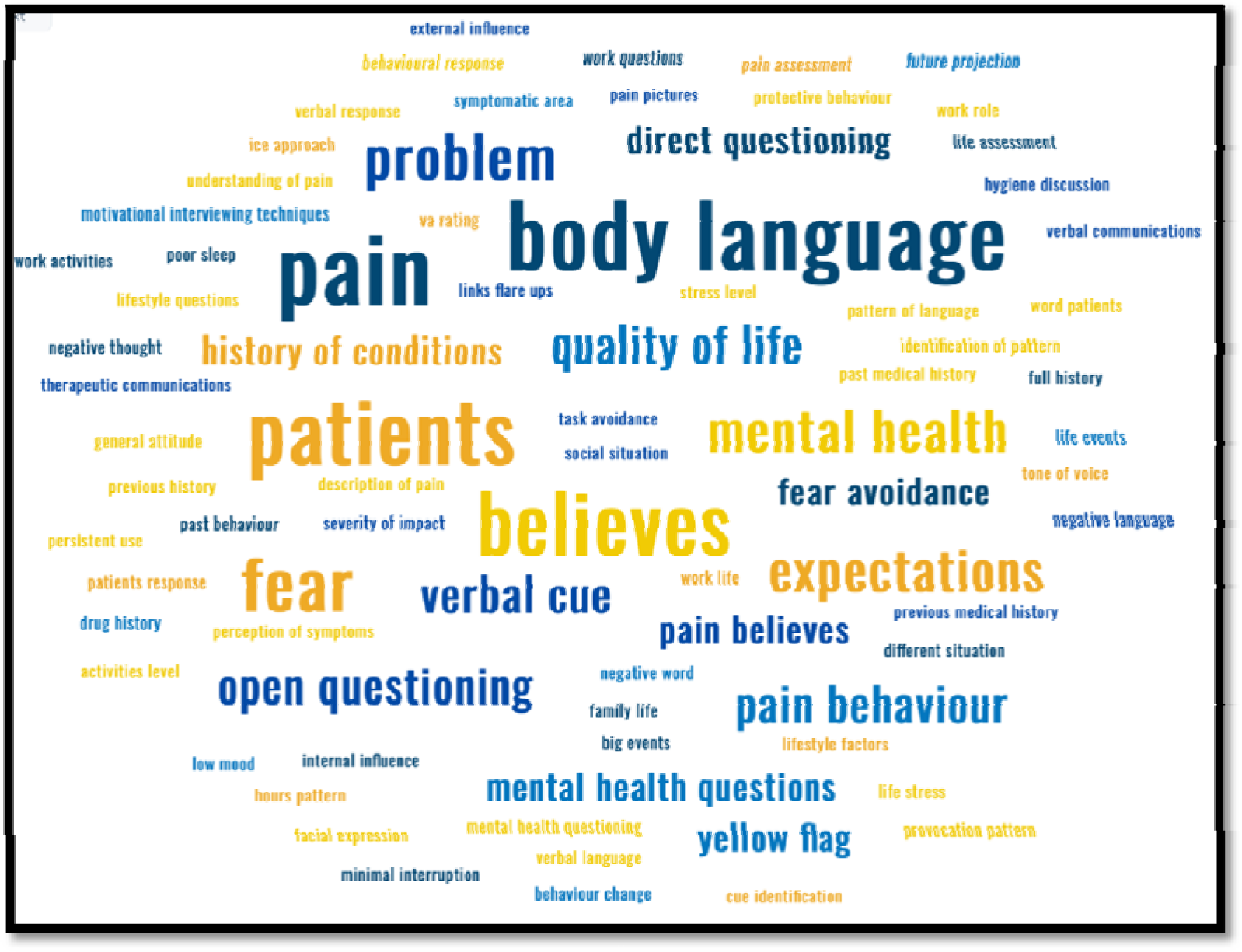
A word cloud depicting ways clinicians explored psychological factors in the subjective assessment.

32 (31.1%) respondents observed their patient’s body language whilst in the subjective assessment. Other respondents asked what their patient’s beliefs (n= 23, 22.3%) and fears (n= 15, 14.5%) were about their injury and what their expectations (n=75, 72.8%) were from the treatment including what the patient understood about the causes and mechanisms of their pain. Pain behaviors were commonly explored (n=25, 24.2%), as was the history of the condition and any mental health history or life events that were recent or relevant. The way in which respondents asked for this information was via open questioning, allowing the patient to tell their story with minimal interruption, listening for patterns, watching for cues and developing a therapeutic relationship to gain trust.

#### Objective Assessment

36 (34.9%) respondents said they assess psychological factors during the objective assessment. Figure 2 shows the word cloud created with respondents’ answers to how they do this.

**Figure 2.**
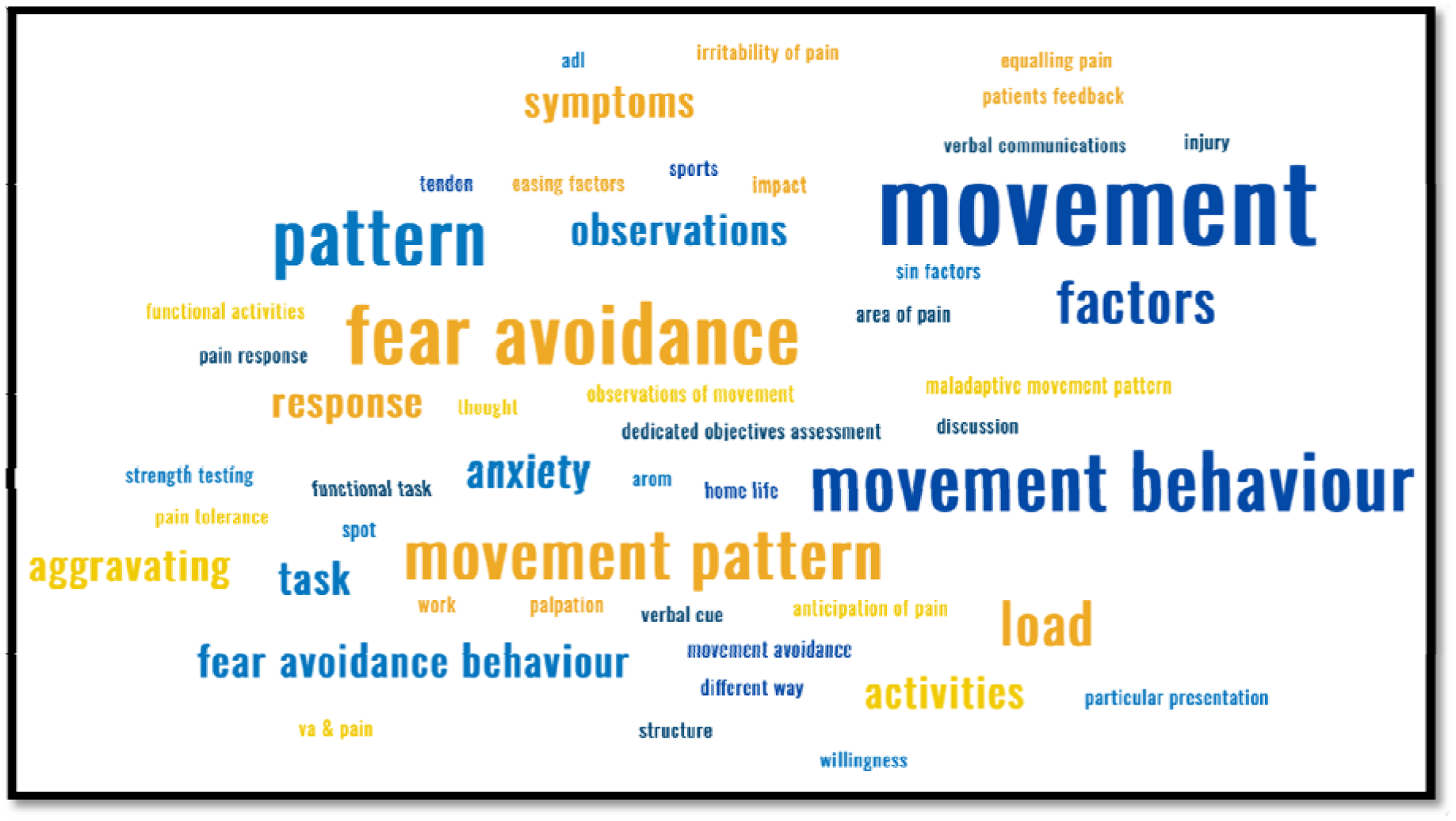
A word cloud depicting ways clinicians explored psychological factors in the objective assessment.

Respondents identified maladaptive movement patterns and fear avoidance behaviours (n=23, 22.3%). Respondents regularly (n=14, 13.6%) identified examining individual’s responses to their pain, the anticipation of pain, willingness to load and the irritability of their pain. Eight (7.8%) of the participants mentioned looking for anxiety towards movement and muscular contraction associated with strength testing. Clinicians consistently reported (n=14, 13.6%) that the signs were often noticed by observing the patient and looking for non-verbal cues, but these cues were not defined.

#### Screening Tools

19 (18.4%) respondents mentioned that they used a specific screening tool to identify mental health or psychological components. 7 respondents used the Tampa Scale of Kinesiophobia (TSK), 3 used the pain catastrophising scale, another 5 used the Örebro Musculoskeletal Pain Screening Questionnaire (OMPQ), [24] 1 each used a pain self-efficacy scale, EQ-5D-5L questionnaire, PH9 + GAD and the MSK HQ measure.

### Assessment of PsychoSOCIAL Components

#### Subjective Assessment

77 (74.8%) respondents assessed psychoSOCIAL components during their subjective assessment alone, whilst 15 (14.6%) combined subjective and objective to determine involvement of PsychoSOCIAL aspects. Some respondents assessed psychoSOCIAL components in the objective examination alone 5 (4.9%).

Some respondents (n=7, 6.8%) said they did not separate psychological from psychoSOCIAL questioning. 23 (22.3%) clinicians used direct questioning about the patient’s lifestyle, 19 (18.4%) asked about their work situation, 23 (22.3%) asked what hobbies and activities their patient’s did and how their pain affected these, 12 (11.6%) asked about their patient’s beliefs, 7 (6.8%) asked what social influences they had, 5 (4.9%) asked what interventions they had tried previously and 3 (2.9%) asked how their sleep was. In the free text section, clinicians reported that they thought it was important to let their patient ‘tell their story’, with little interruptions and using active listening skills.

#### Objective Assessment

25 (24.3%) of the clinicians assessed for psychoSOCIAL factors in their objective assessment. Respondents reported this was done by observing their patient’s Social history, work and socioeconomic status (n=2, 1.9%), behaviours (n=10, 9.7%), how they hold themselves (n=10, 9.7%), their physical literacy (n=5, 4.9%), whether they avoid movements due to fear(n=6, 5.8%), and their motivation towards and completion of their exercises (n=5, 4.9%) and further questioning (n=7, 6.8%). Several of the respondents (n=6, 5.8%) stated they used the same elements as their objective assessment for psychological aspects with examples of fear avoidance behaviours and body language. There was a lack of information around specific traits observable traits that would allow the identification of psychoSOCIAL factors whilst observing alterations in patients’ movements and behaviours. The word cloud in figure 3 amalgamates the subjective and objective screening methods reported by respondents.

**Figure 3.**
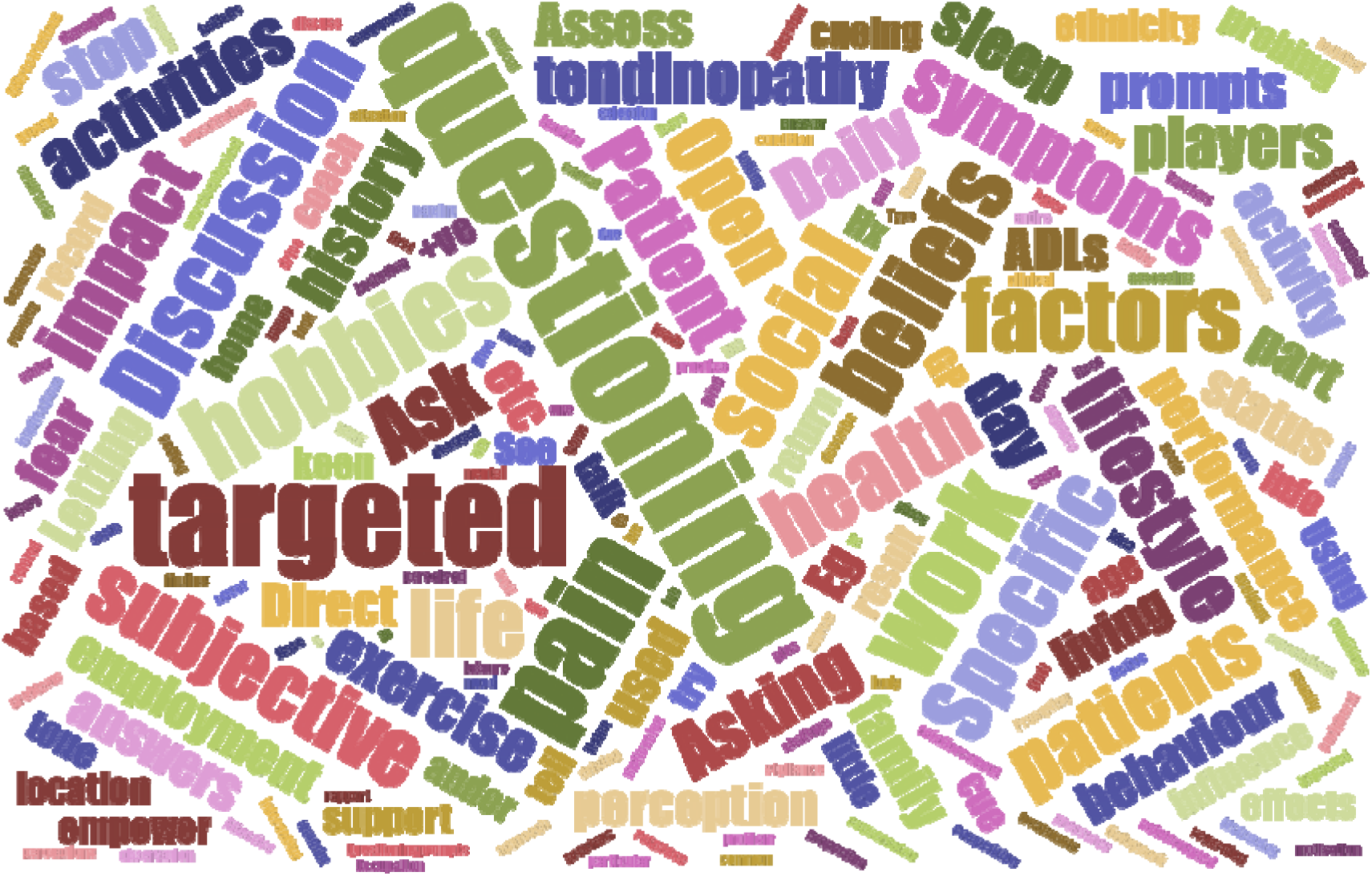
Word cloud depicting the results from respondents’ answers regarding examining psychoSOCIAL components and includes both the subjective and objective elements.

#### Screening Tools

12 (11.6%) of respondents said they chose a specific screening tool to assess psychoSOCIAL factors. The clinicians named the same tools for psychological factors, which were the TSK (n=7, 6.8%) Orebro OMPQ tool (n=5, 4.9%), Pain self efficacy questionnaire (n=3,2.9%). One other identified a series of questions “related to a questionnaire” asking about where individuals work, problems with work colleagues, conflicts with partner/family, or during sports, on the street or driving. No information was given about this questionnaire.

### Prevalence of Constructs in Practice

Respondents were asked to rank the psychological and psychoSOCIAL factors in order of prevalence in their practice.

Table 1 shows the results of which psychological factors clinicians observed most frequently in practice when treating individuals with tendinopathy. Negative pain beliefs, anxiety, maladaptive behaviour, fear of movement, catastrophising behaviour and stress were observed the most frequently. The mean and mode ranks varied by relatively large amounts and Hypervigilance/somatization ranked lower but was not observed as frequently.

**Table 1:** Prevalence of Psychological Factors linked to Tendinopathy in Practice.

| Construct | Mean Rank | Rank range | Mode | No of votes (n/103) |
| --- | --- | --- | --- | --- |
| Negative pain beliefs | 4.79 | 1 to 13 | 1 , 4 & 5 | 97 |
| Anxiety | 4.12 | 1 to 13 | 1 | 93 |
| Maladaptive/Avoidance behaviours | 4.41 | 1 to 12 | 5 | 96 |
| Fear of Movement | 4.40 | 1 to 13 | 3 | 93 |
| Catastrophising Behaviour | 6.26 | 1 to 13 | 8 | 94 |
| Stress | 5.15 | 1 to 12 | 5 & 6 | 91 |
| Self Efficacy | 6.87 | 1 to 12 | 6 | 50 |
| Depression | 7.37 | 1 to 13 | 10 & 11 | 83 |
| Emotional Distress | 7.45 | 1 to 12 | 11 | 82 |
| Hyper-vigilance/Somatisation | 6.46 | 1 to 12 | 3 | 80 |
| Mental Health | 8.37 | 2 to 13 | 6 | 70 |
| Anger | 9.14 | 2 to 12 | 12 | 70 |

Table 2 shows the results of which psychoSOCIAL factors clinicians see the most in clinical practice, ranked in order from 1 to 8 based on number of votes. QOL, work related constructs, sleep quality, education and social interactions were the most common constructs found in practice with the lowest mean ranks and modes.

**Table 2:** Prevalence of PsychoSOCIAL Factors linked to Tendinopathy in Practice.

| Construct | Mean Rank | Rank range | Mode | No of Votes |
| --- | --- | --- | --- | --- |
| Sleep Quality | 3.38 | 1 to 8 | 2 | 88 |
| Work related constructs | 2.92 | 1 to 7 | 1 | 91 |
| Social Interactions/Social Networks and Support | 3.85 | 1 to 7 | 3 | 87 |
| Education/ Health Literacy | 3.54 | 1 to 8 | 3 | 83 |
| Socioeconomic status | 5.05 | 1 to 8 | 6 | 83 |
| Cultural Influences | 5.05 | 1 to 8 | 7 | 78 |
| Quality of Life | 2.74 | 1 to 8 | 1 | 93 |
| Place of residence | 6.69 | 1 to 8 | 8 | 59 |

### Any other psychological or psychoSOCIAL factors?

Clinicians were asked if there were any other constructs they come across in practice. Psychological factors suggested by clinicians include sleep disorders (n=3, 2.9%) and negative beliefs and expectations towards treatment (n=3, 2.9%). Five individuals made further suggestions for psychoSOCIAL factors which included: availability of physiotherapy and other health services in rural areas, participation in high level sport and personality types.

### Most Important Constructs in Practice

Participants were asked to rank from 1 to 5, the most important psychological and psychoSOCIAL factors linked to tendinopathy.

Table 3 shows the results of the 5 most important psychological factors the respondents thought should be considered in practice. Fear of movement was voted the most important, with the mode being 1 and having the greatest number of votes. Other important constructs included mental health, negative pain beliefs, maladaptive behaviours, catastrophising behaviour and anxiety.

**Table 3:**
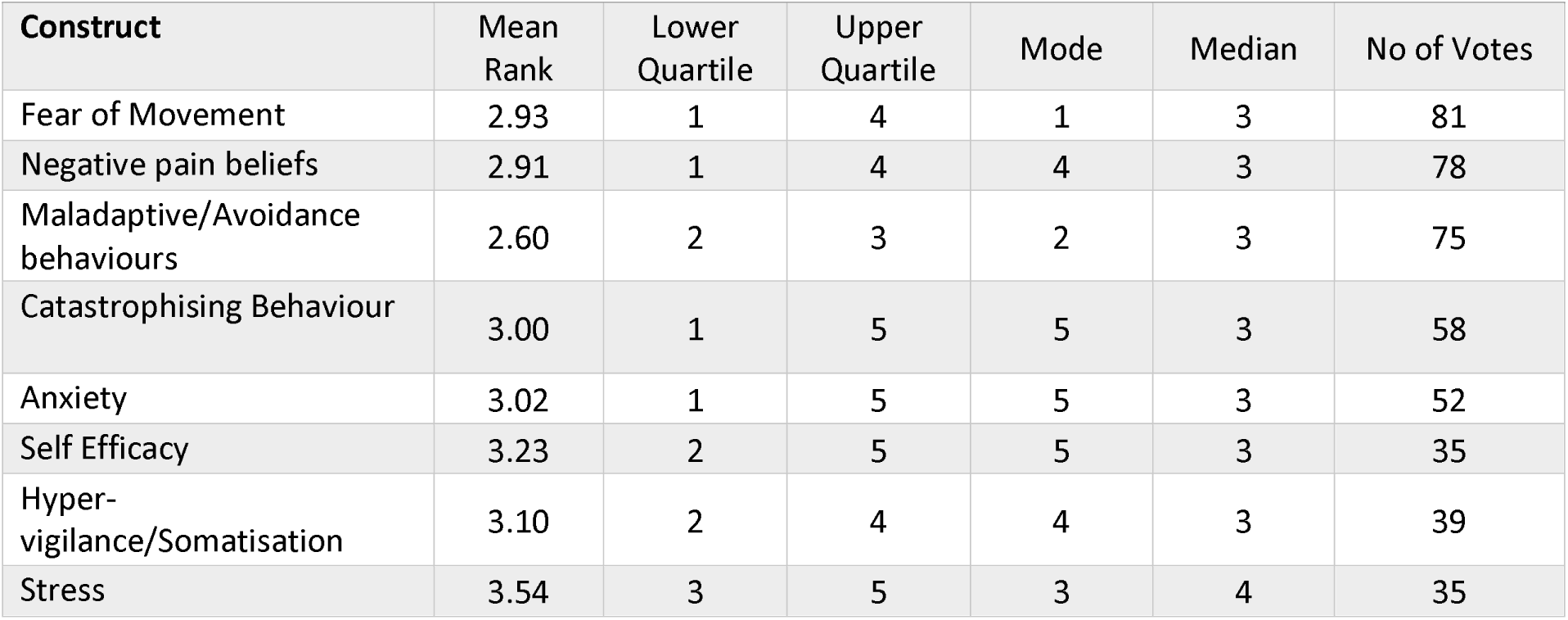
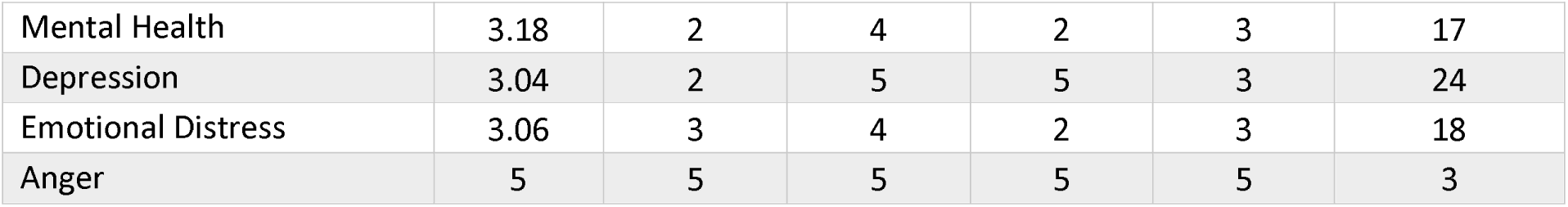
Five most important psychological factors when assessing and treating tendinopathies.

Table 4 shows the respondent’s 5 most important psychoSOCIAL factors when assessing and managing tendinopathy in practice which were QoL, work-related constructs, sleep quality, education/health literacy and social interactions.

**Table 4:** Five most important psychoSOCIAL factors when assessing and managing tendinopathies.

|  |  |  |  |  |  |  |
| --- | --- | --- | --- | --- | --- | --- |
| **Construct** | **Mean Rank** | **Lower Quartile** | **Upper Quartile** | **Mode** | **Median** | **No of votes:** |
| Quality of Life | 2.48 | 1 | 3 | 1 | 2 | 92 |
| Work related constructs | 3.00 | 1 | 4 | 1 | 3 | 90 |
| Sleep Quality | 2.74 | 1 | 4 | 1 | 3 | 85 |
| Education/ Health Literacy | 2.86 | 2 | 4 | 2 | 3 | 87 |
| Social Interactions/Social Networks and Support | 3.21 | 2 | 5 | 3 | 3 | 77 |
| Cultural Influences | 3.51 | 3 | 5 | 5 | 4 | 47 |
| Socioeconomic status | 4.13 | 4 | 5 | 5 | 4 | 31 |
| Place of residence | 3.83 | 2.5 | 5 | 5 | 4.5 | 6 |

### Treatment

The following themes were identified for the types of treatment clinicians offered to target psychological and psychoSOCIAL factors related to a patient’s tendinopathy: education and reassurance, addressing maladaptive behaviours, Cognitive Functional Therapy, behavioural change, referrals and individualised treatment. These are represented in figure 4.

**Figure 4.**
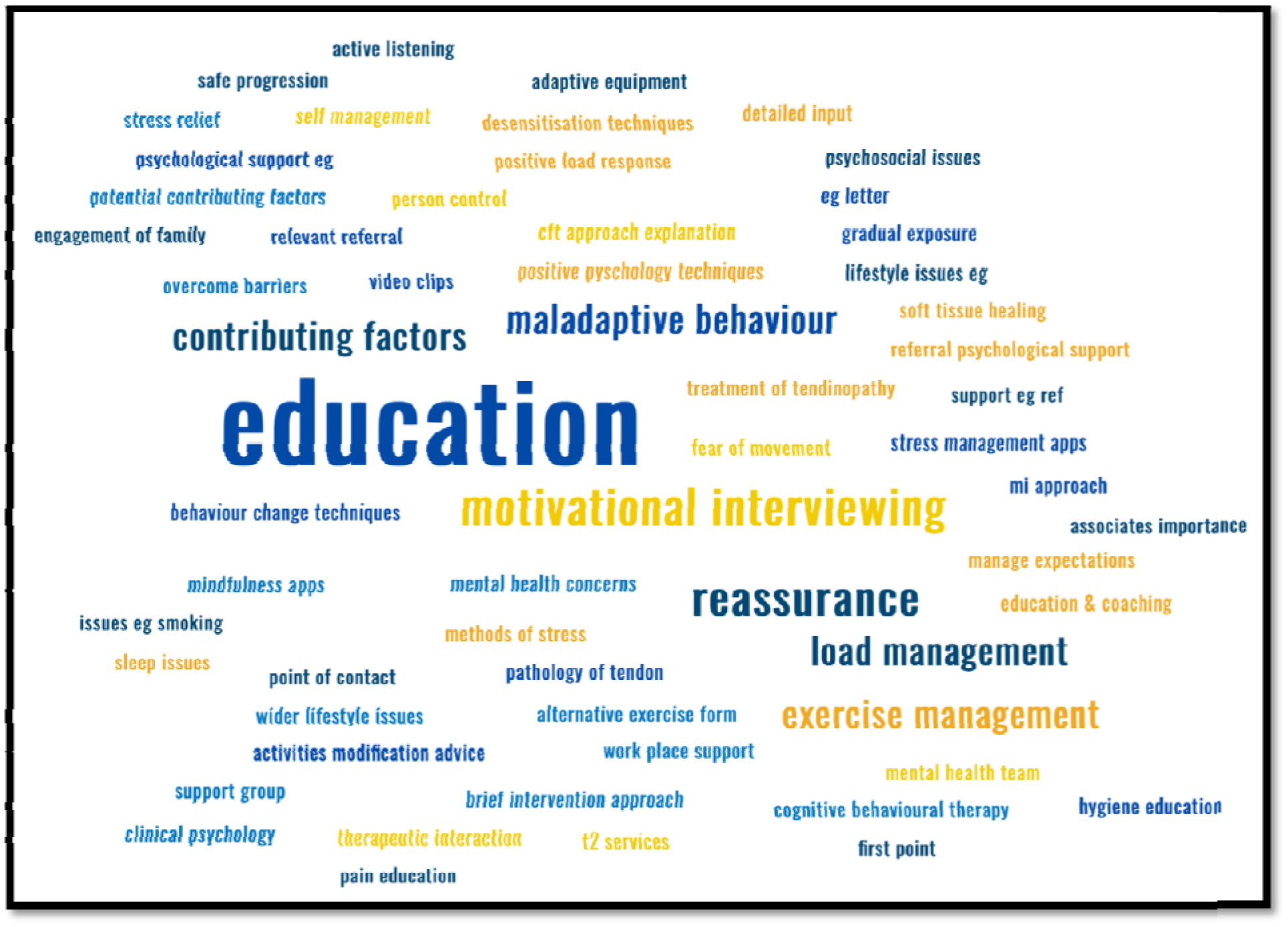
A word cloud representing respondents’ answers about their treatment methods and techniques.

**Figure 5.**
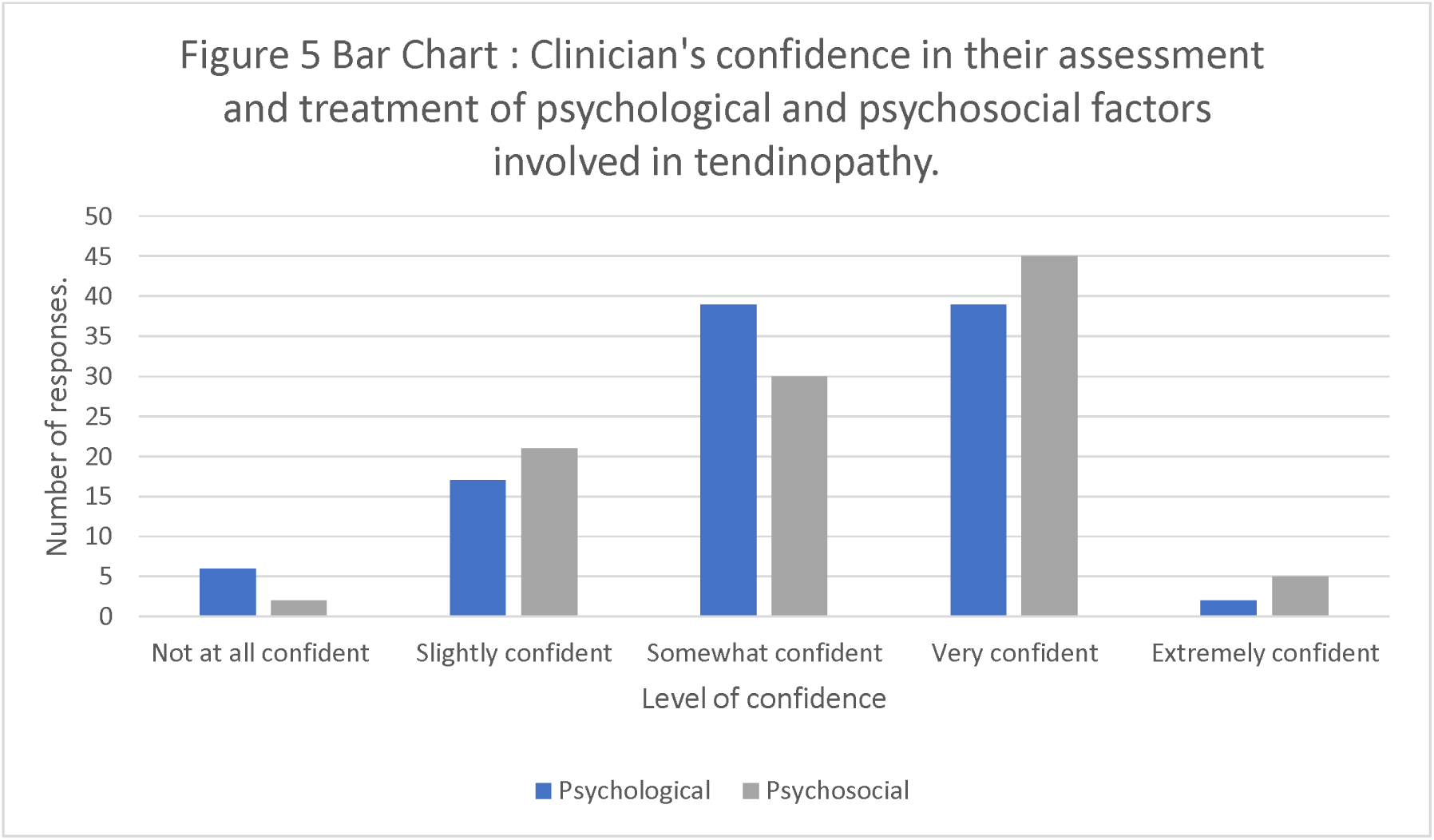
A bar chart identifying clinicians confidence in their assessment and treatment of psychological and psychoSOCIAL factors involved in tendinopathy.

#### Education and Reassurance

68 (66.0%) of the respondents mentioned providing education which included discussing the individual’s beliefs and checking for correct understanding to address catastrophic thoughts. Respondents reported this included reassurance and education around what tendinopathy is and what to expect, and the impact of psychological and psychoSOCIAL factors on recovery and lifestyle changes such as sleep patterns, diet and smoking cessation.

#### Addressing Maladaptive Behaviours

21 (20.4%) respondents mentioned using graded exposure to movements that patients feared and addressing fear and anxiety by using coaching and motivational techniques.

#### Cognitive Functional Therapy/Behavioural Change

6 (5.8%) respondents mentioned using a cognitive functional therapy approach.

#### Referrals

31 (27.9%) clinicians mentioned setting up a referral to mental health teams for further support or towards other resources such as mindfulness or stress management tools, apps, booklets, videos, and support groups.

#### Individualised treatment

9 (30.1%) of the respondents mentioned that the treatment should be tailored to the individual and what matters the most to them by setting goals and highlighting their progress along the way. One answer explained that all treatment options should be presented to the patient so they can make an informed decision.

### Outcome Measures

Respondents were asked whether they use outcome measures to assess psychological and psychoSOCIAL factors. 19 respondents (18.4%) said yes to using outcome measures, whereas 84 (81.5%) respondents said they do not use outcome measures.

Out of the 14 clinicians who answered yes, 7 used the TSK, 5 used the EQ-5D-5L, 4 used a tool for pain catastrophisation or pain self-efficacy, 2 used the Hospital Anxiety Depression Score (HADs). Also mentioned once were the Central Sensitization Inventory (CSI), QOL questionnaires, the Orebro Musculoskeletal pain questionnaire, the Fear-Avoidance Beliefs Questionnaire (FABQ), Patient health questionnaire (PHQ) 9, and Generalised anxiety disorder (GAD) 7 questionnaire.

### Confidence in assessment and treatment

Confidence clinicians felt in their ability to assess and treat psychological and psychoSOCIAL factors is shown. The majority were confident (somewhat confident or very confident or extremely confident) in their ability to assess the psychological (77.9%) and psychoSOCIAL (77.9%) factors. 23 (22.1%) respondents were unconfident (Not at all confident or Slightly confident in their ability to assess either psychological or psychoSOCIAL factors.

### Additional comments

Participants were offered the opportunity to add any further comments regarding their clinical practice. The main themes in the responses were related to time constraints, more integrated care and training needs. Respondents reported that t**ime constraints** meant that they did not have enough time needed to deal with chronic and complex problems and to complete outcome measures. This was particularly reported by those in public health care settings. They reported this effected the effectiveness of their interventions.

Respondents identified that there was a requirement for more **integrated care**, this would ideally involve psychology and counselling support system within departments or teams. This support would provide education and movement therapy, respondents suggested this would facilitate their work and improve patient journeys. T**raining needs** were also highlighted with respondents identifying the belief that undergraduate training needed more prepare the next generation more thoroughly for the demands of managing people with complex psychological and psychoSOCIAL issues.

## Discussion

This is the first study to explore healthcare providers’ knowledge and use of psychological and psychoSOCIAL screening and interventions in the management of patients with tendinopathy. The results show respondents frequently examine these constructs, but formal patient reported outcome measures (PROMs) are seldom utilised. Clinicians’ treatment primarily focuses on education and behaviour change.

Clinicians commonly explore psychological and psychoSOCIAL factors through clinical interactions, relying on subjective and objective examinations rather than formal Patient-Reported Outcome Measures (PROMs) or questionnaires. This contrasts with research literature where formal PROMs are used. [12,13] Instead, clinicians employ open questioning during history-taking, probing into an individual’s story, beliefs, and social situation. Objective assessments involve observing movement patterns and identifying fear avoidance behaviours, aligning with relevant literature.[25] However, the observed lack of formal assessment raises concerns about the accuracy of clinician self-reporting in assessing and treating psychological and psychoSOCIAL factors.

The limited use of formal PROMs is evident, as revealed in a recent service evaluation where only 2% of Achilles tendinopathy patient records included formal questionnaires.[26] Clinicians prefer open questioning and observational assessments during clinical interactions, suggesting a potential need for additional training in formal assessment tools. A significant portion of clinicians (22.1%) lacks confidence in psychological or psychoSOCIAL assessment and treatment. Research emphasises the necessity for training, mentoring, and practice assessment to enhance competency in this area. [27–29] The observed deficiency in formal assessment raises concerns about the accuracy of clinician self-reporting, given their varying competency levels and potential overconfidence. [27,28,30,31] This concern is further compounded when tools are utilised for psychoSOCIAL assessments, as clinicians predominantly resort to psychological patient-reported outcome measures and objective assessments that would normally be considered psychological constructs such as behaviour, fear avoidance and physical literacy. This suggests a potential misinterpretation of psychological and psychoSOCIAL constructs, indicating a need for further training, as highlighted by many respondents expressing a desire for specific training and self-development.

The tool mentioned most frequently was the TSK which is regularly used in the tendinopathy literature [12,13] and can help with identifying sub-groups, specifically in the people with Achilles tendinopathy. [20–22] The most highly ranked variables in both the psychological and psychoSOCIAL domains (fear of movement, negative pain beliefs, maladaptive/avoidance behaviours, catastrophisation and anxiety (psychological constructs) and quality of life, work related constructs, sleep quality, education health literacy and social interactions (psychoSOCIAL constructs) have appropriate validated tools to allow measurement, although these were infrequently reported. Interestingly many of these constructs (fear of movement, fear avoidance beliefs (maladaptive behaviour) and pain beliefs) were recently identified as important for further research based on an international Delphi study with experts and patients. [10] The similarity between the survey results and recently published international Delphi study support the findings and suggest that the identified factors should be used in further research and clinical practice. [10] However, it’s worth noting that factors frequently reported (as seen in tables 1 and 2) didn’t always align with those deemed most crucial (as in tables 3 and 4). This discrepancy may stem from clinicians observing certain factors frequently but not necessarily prioritising them for clinical intervention.

Treatment for identified factors was reported as typically comprising of individualised education, reassurance, addressing of mal-adaptive behaviours and behaviour change. BiopsychoSOCIAL models of care encourage clinicians to explore and manage the patient’s psychological and psychoSOCIAL constructs in tendinopathy clinical care. Respondent bias may exist, especially if those from social media or special interest groups are early adopters or at the forefront of clinical practice. A recent NHS service evaluation found infrequent documentation of psychological or psychoSOCIAL factors in physiotherapists’ notes, suggesting underreporting or recognition without documentation. [26] A notable proportion of respondents opened but did not complete/submit the survey, introducing inherent bias common to survey methodologies.

### Implications for Clinical Practice

Clinicians often assess psychological and psychoSOCIAL factors in tendinopathy assessments but may struggle to identify them effectively due to the infrequent use of validated tools. The findings and recent ICON scoping review recommend considering the use of simple, valid, and reliable Patient-Reported Outcome Measures (PROMs) such as TSK-17, PCS, and HADs. [12,13]These tools serve as useful screening tools and measures of change in relevant factors. Clinicians and researchers should focus on assessing prevalent or highly ranked psychological variables, including fear of movement, negative beliefs, maladaptive/avoidance behaviour, catastrophisation, and anxiety. PsychoSOCIAL variables like quality of life, work-related constructs, sleep quality, education, and health literacy rank highest.

### Conclusion

A high proportion of clinicians’ screen psychological and psychoSOCIAL factors, but few use validated tools due to time constraints and confidence issues. Crucial constructs for consideration include fear of movement, negative pain beliefs, maladaptive/avoidance behaviours, catastrophisation, anxiety, quality of life, work-related constructs, sleep quality, education, health literacy, and social interactions. Future work should explore barriers and facilitators to implementing suitable screening tools and PROMs in clinical practice.

## Data Availability

All data produced in the present study are available upon reasonable request to the authors

